# Awareness of Antimicrobial Resistance and Associated Factors among Poultry Farmers in Osun State, Nigeria: Implications for Surveillance and Stewardship Programs

**DOI:** 10.64898/2026.01.23.26344687

**Authors:** Sunday Charles Adeyemo, Sunday Olakunle Olarewaju, Ifedola Olabisi Faramade, Kehinde Awodele, Eniola Dorcas Olabode, Olujuwon Philip Towoju, Oyelola Eyinade Adeoye, Obehi Are-Daniel, Ayodele Raphael Ajayi, Oladunni Opeyemi

## Abstract

**Background:** Antimicrobial resistance (AMR) is a global public health threat driven significantly by antimicrobial misuse in agriculture, particularly in poultry farming. This study assessed the awareness, knowledge, practices, and associated factors related to antimicrobial resistance among poultry farmers in Osun State, Nigeria.

**Methods:** A cross-sectional study was conducted among 289 poultry farmers selected through stratified random sampling across Osun State. The study included actively practicing poultry farmers aged 18 years and above who used antimicrobials in their operations. Farmers not using antimicrobials were excluded. Data were collected using a pre-tested, structured, interviewer-administered questionnaire and analyzed with SPSS version 27. Descriptive statistics, chi-square tests, and inferential analyses were used to examine relationships between variables.

**Results:** The majority of respondents (89.6%) had heard of AMR, the majority 239 (92.3%) of the respondents heard it from veterinary doctors. The majority (77.2%) also demonstrated good knowledge. Most farmers (89.6%) used antibiotics, with 52.9% using them occasionally. Personal experience (57.8%) was the primary basis for antibiotic selection. About 71.6% implemented biosecurity measures, and 57.8% had received training on AMR. Significant associations were found between knowledge and practice (p<0.001) and between attitude and practice (p<0.001).

**Conclusion:** Despite high awareness, antibiotic misuse persists, driven by factors such as reliance on personal experience and limited veterinary consultation. There is a need for enhanced farmer education, stricter regulatory enforcement, and the implementation of targeted antimicrobial stewardship programs to mitigate AMR risks in poultry farming.

## Introduction

Antimicrobial resistance (AMR) is a formidable global health threat, undermining the efficacy of essential medicines and jeopardizing decades of medical progress [1]. It occurs when bacteria, viruses, fungi, and parasites evolve to withstand antimicrobial treatments, leading to prolonged illness, increased mortality, and soaring healthcare costs [2]. The World Health Organization (WHO) has ranked AMR among the top ten global public health challenges, with projections suggesting it could cause 10 million deaths annually by 2050 if left unchecked [1]. Beyond human health, AMR endangers food security, economic stability, and sustainable development, particularly in low- and middle-income countries where health systems are fragile and surveillance is weak [3].

A primary driver of AMR is the misuse and overuse of antimicrobials in the agricultural sector, especially in livestock production [4]. Poultry farming, in particular, has become a critical hotspot for the emergence and transmission of resistant pathogens due to the intensive use of antibiotics for disease prevention, treatment, and growth promotion [5]. In many regions, antibiotics are administered without veterinary oversight, often at sub-therapeutic doses or for non-therapeutic purposes, accelerating the selection of resistant bacterial strains [2]. These resistant bacteria can spread to humans through direct contact with animals, consumption of contaminated meat and eggs, and environmental pathways such as water and soil contaminated with animal waste [6].

In Nigeria, the largest poultry producer in Africa, the situation is especially concerning with studies indicating alarmingly high rates of antibiotic misuse among farmers, with over 70% using antimicrobials without veterinary prescription [7, 8]. Resistance to commonly used antibiotics such as tetracyclines, fluoroquinolones, and sulfonamides is widespread, and multidrug-resistant pathogens including *Escherichia coli, Salmonella*, and *Campylobacter* are frequently isolated from poultry and farm environments [9]. These trends are exacerbated by weak regulatory enforcement, limited access to veterinary services, low literacy levels, and the pervasive availability of over-the-counter antibiotics [10].

Despite Nigeria’s adoption of a National Action Plan on AMR (2017–2022), implementation at the local level remains inconsistent [15]. Previous studies in Nigeria have focused largely on clinical and microbiological aspects of AMR, with fewer examining the behavioral and socio-cultural dimensions among poultry producers. In Osun State, a key agricultural region in southwestern Nigeria, poultry farming is economically significant, yet there is a dearth of localized data on farmers’ awareness, knowledge, and practices regarding AMR [3]. Understanding these factors is essential for designing effective, context-specific interventions, including surveillance systems and antimicrobial stewardship programs that are feasible and acceptable to farmers.

This study therefore aimed to assess the awareness, knowledge, practices, and associated factors related to AMR among poultry farmers in Osun State. By identifying gaps and barriers to responsible antibiotic use, the findings will inform targeted policies and programs aimed at mitigating AMR risks, safeguarding public health, and ensuring the sustainability of poultry farming in Nigeria.

## Materials and Methods

### Study design and setting

A descriptive cross-sectional study was conducted in Osun State, Nigeria, from July to August 2025. The state comprises 30 local government areas (LGAs) with a significant proportion of the population engaged in poultry farming.

### Study population and sampling

The study included actively practicing poultry farmers aged 18 years and above who used antimicrobials in their operations. Farmers not using antimicrobials were excluded.

In this study, the Leslie Fisher formula for the estimation of sample size in populations greater than 10,000 was used. The formula is expressed as:

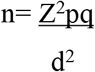

Where

n = required sample size

Z = Standard normal deviate at 95% confidence level = 1.96

p = Proportion of poultry farmers who are aware of AMR, assumed to be 21.4% (0.214) [3] q = Complement of p, calculated as (1 - p) = 0.5

d = Margin of error, which is set at 5% (0.05)

Calculation:

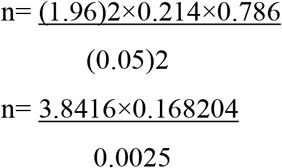

n= 259 respondents.

To cater for possible non-responses, 11.5% is added to the obtained sample size. n= 259 * (1+ 0.115)

n= 259 * 1.115

n= 289

The sample size was 289 respondents

### Sampling technique

A stratified random sampling technique was used for the selection of the sample that best represents the poultry farming landscape of Osun State. Stratification is important in this process because it captures the variation in poultry farming practices across different regions within a state. Stratification Process

Since this study aims to identify the poultry farming zone, the stratification process was made as follows:

1. Poultry Farming Zone Identification: Osun State was divided into geographically distinct regions based on existing administrative divisions (e.g., Osun Central, Osun East, and Osun West). Within each region, major poultry-producing local government areas (LGAs) was identified.
2. Proportional Allocation of Sample Size:

The total sample size, n = 289, was distributed across the selected regions in line with the estimated population of poultry farming in each zone. This means that regions with more concentrations of poultry farms contributed more respondents to the sample while keeping the proportions intact.

3. Random Selection of Respondents within Each Stratum:

Having determined the number of respondents for each region that needed to be selected, poultry farmers were randomly selected within each stratum, which is the LGA. The random selection helps to get rid of bias and presents equal opportunity for all eligible farmers to be represented in the study.

Using stratified random sampling, this research gave a balanced regional representation of the dataset on the awareness of AMR amongst poultry farmers in Osun State. These methods ensured that the findings reflect the true diversity of poultry farming in the state and improve the validity and applicability of the conclusions obtained from the research.

### Data collection

A structured questionnaire was interviewer-administered face-to-face. It covered socio-demographics, awareness and knowledge of AMR, antibiotic use practices, attitudes, barriers to stewardship, and suggestions for improvement. The questionnaire was pretested among 29 poultry farmers (10%) in local governments that were not selected as part of the study.

### Data analysis

Data were analyzed using SPSS version 27. Descriptive statistics summarized socio-demographics and key variables. Chi-square tests examined associations between knowledge, practices, and attitudes. A p-value <0.05 was considered statistically significant.

### Ethical considerations

Ethical approval was obtained from the Health Research Ethics Committee of Osun State University. Informed consent was obtained from all participants, and confidentiality was maintained throughout the study.

## Results

### Socio-demographic characteristics

Most respondents (57.8%) were female, and 26.3% were aged 55 years or older. About 39.4% had tertiary education, and 57.1% operated small-scale farms (<500 birds). Broilers (77.9%) and layers (100%) were the most reared birds (Table 1).

**Table 1.** Socio-demographic characteristics of respondents (N=289)

| **Variables** | **Frequency** | **Percentage** |
| --- | --- | --- |
| **Age (in years)** |  |  |
| 18- 25 | 46 | 15.9 |
| 25-34 | 54 | 18.7 |
| 35-44 | 60 | 20.8 |
| 45-54 | 53 | 18.3 |
| 55 years above | 76 | 26.3 |
| **Gender** |  |  |
| Male | 122 | 42.2 |
| Female | 167 | 57.8 |
| **Level of Education** |  |  |
| No formal education | 30 | 10.4 |
| Primary | 85 | 29.4 |
| Secondary | 60 | 20.8 |
| Tertiary | 114 | 39.4 |
| **Duration of poultry farming** |  |  |
| < 1 year | 43 | 14.9 |
| 1-5 years | 91 | 31.5 |
| 6-10 years | 103 | 35.6 |
| > 10 years | 52 | 18 |
| **Size of poultry farm** |  |  |
| Small scale | 165 | 57.1 |
| Medium scale | 97 | 33.6 |
| Large scale | 27 | 9.3 |
| **Types of poultry (Multiple choice)** |  |  |
| Broilers | 225 | 77.9 |
| Cockerels | 49 | 17 |
| Layers | 289 | 100 |
| **Farm location** |  |  |
| Osogbo | 80 | 27.7 |
| Ikirun | 61 | 21.1 |
| Ile Ife | 39 | 13.5 |
| Osu | 17 | 5.9 |
| Ilesha | 45 | 15.6 |
| Ifon | 33 | 11.4 |
| Okinni | 14 | 4.8 |
| **Farm sales target** |  |  |
| Meat | 61 | 21.1 |
| Eggs | 48 | 16.6 |
| Both | 180 | 62.3 |

### Awareness and knowledge of AMR

Findings revealed that majority of the respondents 259 (89.6%) have heard of antimicrobial resistance and the majority 239 (92.3%) of the respondents heard it from veterinary doctors.

Two hundred and twenty-three (77.2%) respondents are aware that the inappropriate use of antibiotics in poultry can lead to AMR. Most respondents 178 (61.6%) are aware that resistant infections in humans can come from eating or handling poultry with antibiotics residue.

Also, 233 (77.2%) respondents know that using antibiotics too frequently, using antibiotics without prescription, not completing the full course of antibiotics and giving animals antibiotics that were meant for human consumptions are the causes of AMR while 66 (22.8%) respondents don’t know the causes of AMR.

The majority of the respondents 223 (77.2%) believed that following veterinary instructions when using antibiotics reduces the risk of resistance and 253 (87.5%) believes that antibiotics have side effects. Overall, 233 (77.2%) respondents had good knowledge of AMR (Figure 1).

**Figure 1.**
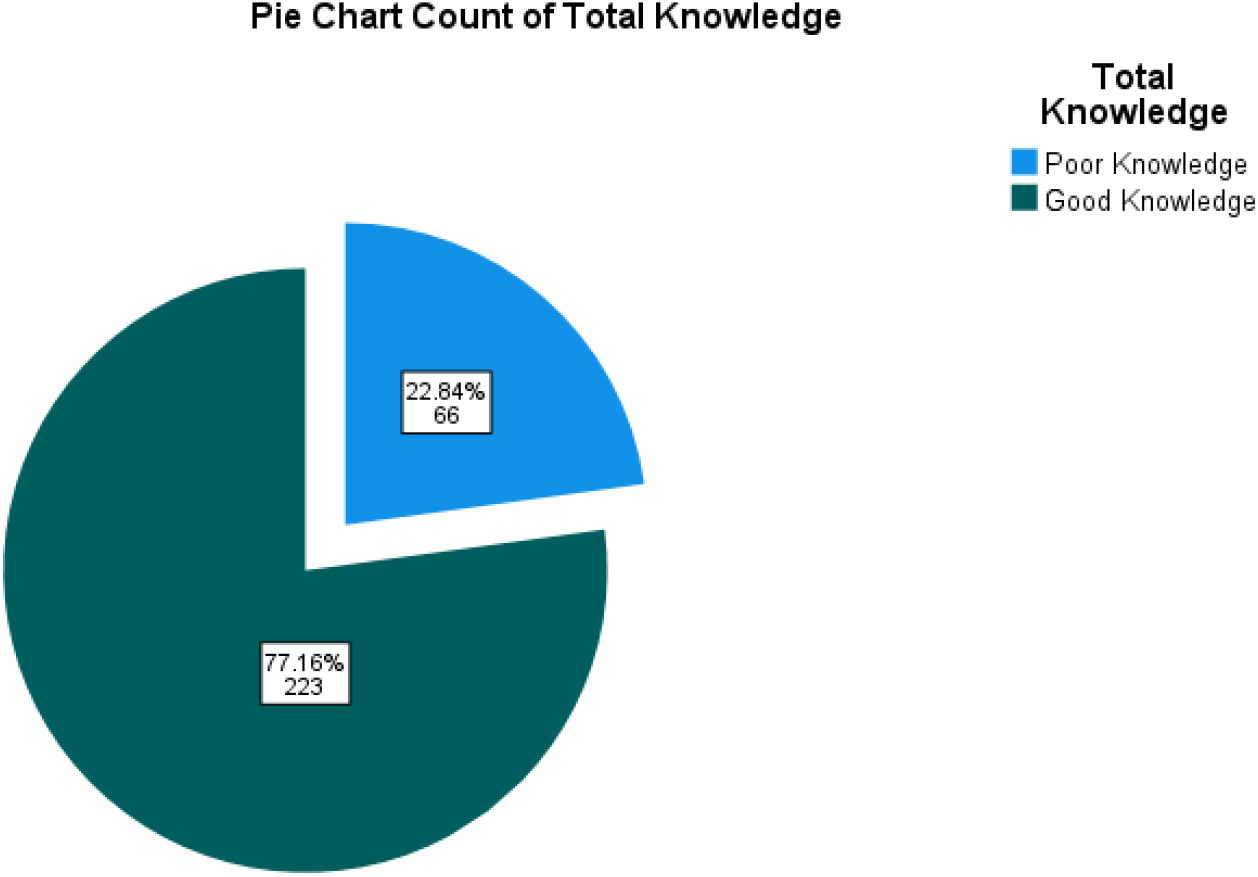
Overall knowledge of Antimicrobial resistance.

### Practices related to antimicrobial use

Most farmers (89.6%) used antibiotics, with 52.9% using them occasionally. The primary reason for use was treatment of sick birds (36%). Only 28.7% followed veterinary prescriptions, while 57.8% relied on personal experience (Table 2). Overall, majority of the respondents 259 (89.62%) have a good level of practice while few respondents 30 (10.38%) have poor level of practice.

**Table 2:** Practices related to antimicrobial use (N=289)

| **Variables** | **Frequency** | **Percentage** |
| --- | --- | --- |
| **Do you use antibiotics in your poultry farm?** |  |  |
| Yes | 259 | 89.6 |
| No | 30 | 10.4 |
| **If yes, how often do you use antibiotics?** |  |  |
| Weekly | 35 | 12.1 |
| Monthly | 28 | 9.7 |
| Occasionally | 153 | 52.9 |
| Only if there is health issue | 37 | 12.8 |
| Only when prescribed by the veterinarian | 36 | 12.5 |
| **Why do you use antibiotics?** |  |  |
| I don't use | 30 | 10.4 |
| To treat sick birds | 104 | 36 |
| To prevent disease outbreak | 47 | 16.3 |
| To promote faster growth | 40 | 13.8 |
| On the advice of a veterinarian | 68 | 23.5 |
| **Is it necessary to consult a veterinarian?** |  |  |
| Yes | 239 | 82.7 |
| No | 50 | 17.3 |
| **How do you decide on antibiotics to use and when to use them?** |  |  |
| Personal experience | 167 | 57.8 |
| Prescription from a veterinarian | 83 | 28.7 |
| Advice from fellow farmers | 9 | 3.1 |
| Others | 30 | 10.4 |
| **Do you consult a veterinarian only after attempted treatment on sick birds have not yielded a positive response?** |  |  |
| Yes | 96 | 33.2 |
| No | 193 | 66.8 |

### Attitudes and barriers to antimicrobial stewardship

The majority (57.8%) considered reducing antibiotic use very important, and 71.6% implemented biosecurity measures. Challenges included bird death (47.8%), drug cost (13.1%), and bird resistance (15.6%). About 57.8% had received AMR-related training (Table 3). Overall, majority of the respondents 244 (84.43%) have good attitude while few respondents 45 (15.57%) have poor attitude towards antimicrobial stewardship.

**Table 3:** Attitudes and barriers to antimicrobial stewardship (N=289)

| Variable | Frequency | Percentage |
| --- | --- | --- |
| **How important to you is it to reduce the use of antibiotics in poultry farming** |  |  |
| Not Important | 45 | 15.6 |
| Slightly Important | 77 | 26.6 |
| Very Important | 167 | 57.8 |
| **Do you implement any biosecurity measures to prevent disease outbreaks on your farm** |  |  |
| Yes | 207 | 71.6 |
| No | 82 | 28.4 |
| **In your opinion, what are the dangers of misuse or overuse of antibiotics in Poultry** |  |  |
| Poor growth rate in poultry | 228 | 90.8 |
| Antibiotics residue in meats or eggs | 228 | 90.8 |
| Development of resistant infection in humans and animals | 221 | 88 |
| Economic losses | 192 | 76.5 |
| **What challenges do you face in using antibiotics responsibly in poultry farming** |  |  |
| Birds resistance | 45 | 15.6 |
| Death | 138 | 47.8 |
| Fake and expensive drugs | 38 | 13.1 |
| None | 68 | 23.5 |
| **Have you ever received training on antimicrobial use or resistance** |  |  |
| Yes | 167 | 57.8 |
| No | 122 | 42.2 |
| **If no, would you be interested in such training programs** |  |  |
| Yes | 152 | 52.6 |
| No | 147 | 47.4 |

### Relationship between knowledge, attitude towards stewardship and practices related to antimicrobial use

A significant association was found between knowledge and practice (χ^2^=100.435, p<0.001), with 86.4% of those with good knowledge demonstrating good practice. Similarly, a significant relationship existed between attitude and practice (χ^2^=115.970, p<0.001), where 81.1% with positive attitudes had good practices.

## Discussion

This research offers perspectives on the knowledge, awareness and behavior pertaining to antimicrobial resistance among the poultry farmers in Osun State Nigeria. The results confirm a paradox: though a large percentage of farmers (89.6%) stated that they were aware of AMR and had good knowledge (77.2%), there are still considerable discrepancies between knowledge and practice. This contradiction emphasizes the important interaction between socio-economic, educational, and systemic factors impacting the antibiotic use in agricultural environments [16].

The awareness level is high in the current study compared with previous studies in sub-Saharan African where the study found low awareness level among the farmers [17, 18]. This change could be an indication of a rise in the use of the veterinary services, media campaigns or involvement in agricultural extension programs over the last few years [5]. It is important to note that veterinarians were mentioned as the most important source of AMR information and emphasized as educators and gatekeepers in antimicrobial stewardship [22]. Nevertheless, this knowledge does not eliminate the fact that the use of personal experience (57.8%) as opposed to the advice of a professional to select and administer antibiotics suggests that there is still a strong adherence to informal knowledge systems [3]. This has been witnessed in other low and middle income (LMIC) countries in which farmers might sacrifice experience learning and formal veterinary advice in favor of short-term economic returns in case of limited access to professional services at low costs [7].

The trends of antibiotic use reported in this study are in line with the larger trend in the country of Nigeria and more generally on other areas where antibiotics are commonly employed as a growth promoter or prophylaxis and not in actual treatment [8, 10]. The occasional usage as noted by more than half of the farmers implies that they have reactive but not preventive attitude to the health of their animal which could be due to poor biosecurity, high disease burden, or limited financial means to invest in non-antibiotic options [6]. Regarding the use of biosecurity, the report is encouraging as more farmers (71.6%) acknowledged adopting good practices, a positive move towards decreasing the number of cases and the use of antibiotics. Nevertheless, the fact that some issues like the death of birds, fake medicines, and expensive veterinary services still persist, means that structural and economic constraints continue to be a major barrier to the responsible use of antibiotics [19].

The excellent statistical correlations between knowledge and practice and the attitude and practice support the significance of combined teaching interventions where a deficit in information is not the only cause of practice failure but also motivational and contextual obstacles [3]. Although one requires knowledge, it is not enough to modify the behavior without accompanying transformations in the access to veterinary services, affordable options, and supportive policies. The fact that the farmers who held positive views concerning antibiotic reduction were more responsive to good practices implies that interventions based on the idea of economic gain, i.e. lowered drug expenses and enhanced availability of the so-called antibiotic-free products, may be more effectively received than those that emphasize only on the potential dangers to the health of the populace [20].

More so, the study points to an increased necessity of improved surveillance and monitoring systems, specific to the Nigerian context. The existing AMR surveillance is still fragmented and under-resourced, which does not allow monitoring the trends of resistance and measuring the effects of interventions [21]. The development of laboratory capacity, harmonization of data collection, and One Health strategies that connect human, animal, and environmental health sectors are all necessary to a coordinated reaction to AMR [22].

There are some limitations of this research that need to be noted. Cross-sectional design does not allow the use of causal inferences and self-reported information can be affected by social desirability bias. Longitudinal designs, mixed-methods, and direct observation to on-farm practices should be considered in the future in order to gain further insights into the problems behind the use and resistance to antibiotics.

## Conclusion

The present study reveals that, although poultry farmers in the Osun State are well informed on AMR, it is not enough to educate them in order to bring transformative change in their practices concerning the use of antibiotics. It requires a multi-pronged approach, which will involve educating the farmers and increasing the availability of veterinary services, the implementation of laws regulating the sale of antibiotics, the encouragement of alternatives, and the presence of efficient surveillance systems. With solutions to the systemic and behavioral forces of AMR, Nigeria will be in a better position of attaining sustainable poultry production and protecting the health of animals and people in the future.

## Acknowledgments

The authors thank the poultry farmers of Osun State for their participation and the Department of Public Health, Osun State University, for institutional support.

## Ethical Approval

Ethical approval was granted by the Health Research Ethics Committee of Osun State University.. Informed consent was obtained from all participants. The ethical principles and guidelines set out by the Declaration of Helsinki, the Belmont Report, and other relevant documents were followed during the conduct of the study.

## Consent for publication

Not applicable

## Clinical trial

Not applicable

## Availability of data and material

The data for this study is provided within the manuscript

## Author’s contribution

SCA, SOO, and IOF worked on the study design; SOO, KA, and OPT collected data; SOO, OEA and SCA supervised the project; SCA, EDO, OA and ARA analyzed the data while SOO, OO and SCA ensured ethical compliance. SOO, EDO and SCA were major contributors in writing the manuscript. All authors read and approved the final manuscript.

## Funding

The authors did not receive any funding for the study.

## Competing interests

The authors know no competing interest for this study.

## Notes

### Competing Interest Statement

The authors have declared no competing interest.

### Funding Statement

The author(s) received no specific funding for this work.

### Author Declarations

Ethical approval was granted by Health Research Ethics Committee of Osun State University with approval number- UNIOSUNHREC2025/PBH/003

